# Umbilical Cord Care among Women 15-49 Years in Uganda; a National Cross Sectional Study

**DOI:** 10.1101/2025.08.13.25333574

**Authors:** Nasser Kasadha, Gertrude Namazzi, Monica Okuga, Ronald Wasswa, Rornald Kananura, Darius Kajjo, Joaniter Nanakabirwa, Peter waiswa

## Abstract

**Background:** Globally, 203000 children die of neonatal sepsis annually. Neonatal Sepsis, partly caused by poor umbilical cord hygiene, is responsible for 18.2% of neonatal deaths in Uganda. While the national newborn care includes cord care practices, the extent of compliance with these recommendations in Uganda remains unclear.

**Objective:** This study aimed at estimating the prevalence of applying non-recommended substances for umbilical cord care, identify substances used, and assess factors associated with use of non-recommended substances.

**Methods:** A secondary data analysis was conducted among 19835 women 15-49 years using data from the 2023 Uganda Situation Analysis of Newborn health. Women were included if they had a live birth within the past one year prior to the survey. Data on substances used for cord care, socio-demographic factors, maternal factors, and healthcare service utilization was abstracted using a pretested data abstraction tool. Modified Poisson models from generalized linear models family with log link function and clustered standard errors were used to determine factors associated with use of non-recommended substances for umbilical cord care, using Stata version 15.

**Results:** The prevalence of using non-recommended substances for umbilical cord care was 14%. Some of the non-recommended substances used in this context included body ointments, body powder, ash and various types of oils. The analysis revealed that maternal age 15-19 (aPR=2.2, 95% CI: 1.1-4.5), religion; protestant (aPR=2.3, 95% CI: 1.2-4.2) and Muslim (aPR=0.5, 95% CI: 0.4-0.7), not attending postnatal care (aPR=0.4, 95% CI: 0.2-0.7), were significantly associated with using non-recommended substances for umbilical cord care.

**Conclusion:** The prevalence of applying non-recommended substances for umbilical cord care in Uganda was moderately high. Enhancing maternal education on proper cord care during postnatal care visits could reduce the use of non-recommended substances for umbilical cord care. Educating teenage mothers about recommendable cord care should be prioritized during postnatal care sessions.

## Introduction

The first 28 days of life represent the most vulnerable period for a newborn, characterized by significant physiological transitions and high risk of morbidity and mortality. Globally, an estimated 2.3 million neonates died in 2022, translating to nearly 6,400 deaths everyday (1). According to United Nations Children’s Fund (UNICEF) 2021 neonatal mortality (NM) report, sub-Saharan Africa accounted for over 1.2 million neonatal deaths annually, with an average neonatal mortality rate (NMR) of 27 deaths per 1000 live births (1, 2). In Uganda, NMR remains high, at 22 per 1,000 live births, contributing to 42% of all under-five deaths (3).

The majority of deaths in Uganda are attributed to preventable causes including sepsis, tetanus, diarrhea, prematurity, and birth asphyxia (4). Among these, neonatal sepsis, often linked to inadequate umbilical cord care, accounts for approximately 15% of neonatal deaths (5, 6). Furthermore, suboptimal umbilical cord care related sepsis is implicated in approximately 700,000 neonatal deaths per year globally (7, 8, 9). Although institutional delivery and antenatal care attendance have increased in the past decades from 36% in 2001 to 86% in 2022 and 93% in 2001 to 99% in 2022 respectively, it does not resonate with NM reduction trends, for reasons related to little or no marked improvement in community based newborn care practices including umbilical cord care (3, 10).

Evidence based recommendations for umbilical cord care varies by setting. In high resource environments, dry cord care is advised, while in low resource settings or where home births are more prevalent, 4 % chlohexidine solution or gel is the recommended standard (11, 12), and other substances other than 4% chlorhexidine solution or gel are regarded non-recommended. Despite strong evidence supporting it’s use, including demonstrated reduction in NM in south Asia and other regions, adherence to these guidelines remains suboptimal in many low and middle income countries (13, 14, 15).

More efforts have been exerted to improve neonatal outcomes including post natal care checks within 24 hours, on day 3 and between 7-14 days, and the more recent recommendation of up to 8 ANC visits to increase contact of mothers with skilled providers (12). However, in Uganda and similar settings, studies have shown widespread use of non-recommended substances such as cow ghee, breast milk, Vaseline, ash, cooking oil, and traditional herbal mixtures for umbilical cord care, that have been associated with neonatal sepsis (8, 16).

This was revealed in one study in western Uganda where 70% of mothers used harmful substances that resulted in cord infections (9). Contributing factors included; maternal age, primiparity and home delivery. Similar patterns have been observed in Ethiopia and elsewhere, where cultural practices drive the use of these substances under the belief that they promote healing, and prevent infections and/or bleeding and hasten cord drying (8, 17).

The use of non-recommended substances is closely associated with poverty, low or no education, young motherhood, home birth deliveries, limited access to healthcare, reliance to traditional birth attendants (TBAs) and lack of antenatal care (ANC) attendance (7, 8, 9, 16, 18). Structural and systemic constraints such as low ANC coverage, delayed clinical guidelines updates, inadequate postnatal care attendance, and inconsistent availability of chlorhexidine further exacerbate the problem (3, 8, 9, 12, 19, 20, 21).

The World Health Organization (WHO) emphasizes hygienic cord care as a core component of essential newborn care, recognizing it’s role in reducing neonatal infections and mortality (22). However, in many low and middle income countries including Uganda, uptake remains limited due cultural beliefs, misinformation, socio-demographic factors, and health system constraints (17, 23).

Suboptimal umbilical cord care remains a critical contributor to neonatal sepsis and preventable neonatal mortality in Uganda (9, 24), especially among families from the lower socio-economic backgrounds, which often lack access to skilled birth attendants, clean delivery and living environments, and essential newborn care supplies, factors that increase the reliance on harmful practices including use of non-recommended substances for cord care exposing newborns to otherwise preventable infections (25, 26). This inequity exposes the most vulnerable neonates to life threatening infections, significantly undermining national efforts to reduce newborn deaths.

While the United Nations (UN) has set a global target of reducing NM to fewer than 12 deaths per 1,000 live births by 2030 under the Sustainable Development Goal (SDG) 3, subtheme 3.2, Uganda’s national target is to reduce neonatal deaths from 27 per 1000 live births to 19 per 1000 live births by 2025 (27, 28). Although some progress has been made, with a neonatal mortality declining from 27/1000 live births in 2011 to 22 in 2022 (3), this remains insufficient. Uganda would need to move faster by accelerating it’s rate of reduction to approximately 2 deaths per 1000 live birth per year to remain on truck for the global 2030 agenda. This slow progress reflects persistent gaps in equitable access to quality maternal and newborn care, especially among disadvantaged populations (29). Without targeted community level interventions to eliminate harmful practices like poor umbilical cord care, these gains risk stalling.

Failure to urgently address NM, including deaths resulting from suboptimal cord care poses a significant threat to Uganda’s ability to achieve it’s national and global child survival targets. Neonatal deaths currently account for approximately 61% of all infant deaths and 42% of under-five mortality (3, 27), making it the leading cause of child mortality in the country. If unaddressed, these preventable deaths are likely to undermine Uganda’s efforts to reduce projected infant mortality from 41 to 34 per 1,000 live births, and under-five mortality rate from 62.2 to 30 per 1,000 live births by 2025 (27).

Beyond the immediate human cost, persistently high neonatal and child mortality rates have broader implications for Uganda’s life expectancy, which remains lower than the global average (3, 30). Because these deaths occur early in life, they disproportionally reduce national life expectancy at birth, distorting long term health indicators and masking progress in other population segments (30, 31, 32). Moreover, frequent early childhood deaths represents a direct loss of future human capital and productivity, thereby weakening the labor force, slowing economic growth, and straining health and social systems (33).

Although the 2022 Uganda Essential Maternal and Newborn Clinical Care Guidelines (EMNCC) address newborn health, including infection prevention, they fall short of explicitly recommending the use of 4% chlorhexidine solution or gel for umbilical cord care (12, 34). Additionally, the guidelines lack consistent messages on cord care during critical periods such as antenatal and postnatal care visits, where maternal education on newborn health is most impactful. This noted silence in these guidelines is presumed to stem, in part from lack of nationally representative data on prevalence and partners of using non-recommended substances for cord care in Uganda.

Therefore, this study aims to fill the evidence gap by determining the prevalence of non-recommended cord care practices, identifying the specific substances used, and exploring factors associated with their use among women aged 15-45 years in Uganda. Addressing NM through low-cost interventions such as recommended umbilical cord care is thus not only a public health priority but also a strategic investment in Uganda’s human development and long term prosperity.

## Methods

### Study design and setting

This retrospective cross-sectional study utilized secondary data accessed on 10/05/2024 from the 2023 Uganda Newborn Situational Analysis survey (SITAN), conducted national wide. The SITAN comprised of three components: a secondary data analysis including a scoping literature review, a hospital readiness assessment, and a community-based survey across all 15 Uganda Demography and Health Survey sub-regions. The survey evaluated newborn care practices, mortality, barriers and delays in care and referral pathways. The community based survey, which formed the basis of this study involved a weighted sample of 19835 women of reproductive age and assessed the capacity of community-level health systems to provide newborn care, including community sensitization, continuity of care, emergency referrals, and newborn health practices. Detailed SITAN methodology is available elsewhere (35).

### Population and Sample Size

This study recruited a weighted sample of 19835 women 15-49 years from a nationally representative 2023 Uganda newborn situation analysis survey. Eligible participants were women with a history of a live birth within one year preceding the survey. All participants meeting this criterion were included in the final analysis except those with missing data on key study variables (ANC, PNC, mode and place of delivery and the outcome variable. This large, population based sample ensured sufficient statistical power and generalizability of findings to the wider Ugandan population. By including the full sample from the parent dataset and excluding only incomplete records, the study maximized internal validity while minimizing selection bias.

Data was abstracted using a pretested data abstraction tool by the principle investigator with help of two trained research assistants.

### Study variables and measurements

The study’s dependent variable was application of any substance for umbilical cord care, and substances categorized into recommended and non-recommended substances.

Non-recommended care was defined as application of substance other than 4% chlorhexidine solution or gel and normal saline for healthcare facility based cord cleansing. The prevalence of using non-recommended care substances was calculated as number of women who used non-recommended substances other than 4% chlorhexidine solution or gel divided by the total sample size, and presented as percentage with its 95% CI.

The independent variables included: Age of the mother; categorized into four groups (15-19, 20-24, 25-29 and 30 years and above) based on Uganda demographic and health survey classification and sample size (3); Maternal education: categorized into three groups (none/primary, secondary and tertiary/university); Place of residence (urban and rural); Employment status of the mother: (employed and un-employed); Religion: categorized into five groups (Anglican, Catholics, Muslims, Adventists and Pentecostals); Mode of delivery: (spontaneous vaginal delivery and cesarean section); ANC: (less than 4 visits and 4 and above visits); Maternal PNC attendance:(attended and those who did not attend), and place of delivery categorized into two groups (health facility and home/TBA). The choice of these variables was informed by the conceptual framework in the introduction section.

Maternal age, place of residence (rural or urban), religion, and mode of delivery, maternal education, ANC and PNC attendance were measured on a nominal scale and maternal education measured on an ordinal scale.

### Statistical Analysis

Data was managed using Stata version 15. Percentages and frequencies were used to summarize variables. Modified Poisson bivariate analysis was conducted to select variables to be included at multivariate analysis. All variables with P.values <0.2 were considered significant at bivariate and, therefore included in multivariate models.

Modified Poisson models from generalized linear models (GLM) family with log link function, reporting clustered standard errors were the modeling approach. Only variables with p.values <0.2 at bivariate were considered for multivariate analysis.

Only variables with P-value < 0.05 were considered significantly associated with use of non-recommended substances for cord care.

Significance of interaction was checked manually to check for possible interaction between independent variables. Adjusted prevalence ratios with their 95% confidence intervals and p-values were reported.

## Results

This study was set out to determine the prevalence of using non-recommended substances for cord care, substances used and factors associated with their use. Out of 19835, 6,463 (32.6%) of the mothers were 20-24 years old, and 5477 (27.6%) Catholics. Majority of the participants had no education or had acquired primary education 10440 (52.6%), 12300 (62.0%) were urban dwellers and 12760 (64.3%) employed. Majority, 13999 (70.6%) of the respondents attended four and above ANC sessions during their last pregnancy, and most (87.9%) of these delivered from healthcare facilities **(Table 1).**

**Table 1.** Socio-demographics and obstetrics characteristics of respondents.

| **Variables** | **Frequency(n)** | **Percentage** |
| --- | --- | --- |
| **Age** |  |  |
| 15-19 | 4179 | 21.1 |
| 20-24 | 6456 | 32.5 |
| 25-29 | 4430 | 22.3 |
| 30 and above | 4770 | 24.0 |
| **Religion** |  |  |
| Anglicans | 4764 | 24.0 |
| Catholics | 5504 | 27.8 |
| Muslims | 3943 | 19.9 |
| Adventists | 966 | 4.9 |
| Pentecostal | 4658 | 23.4 |
| **Level of education** |  |  |
| None or primary | 10469 | 52.8 |
| Secondary | 8220 | 41.4 |
| Tertiary/University | 1146 | 5.8 |
| **Place of residence** |  |  |
| Urban | 12,300 | 62 |
| Rural | 7,582 | 38 |
| **Employment status** |  |  |
| Employed | 9547 | 48.1 |
| Unemployed | 10288 | 51.9 |
| **ANC Visits** |  |  |
| <4 Visits | 5836 | 29.4 |
| 4 and above visits | 13999 | 70.6 |
| **Maternal PNC Visits** |  |  |
| Yes | 12281 | 62 |
| No | 7554 | 38 |
| **Mode of delivery** |  |  |
| Spontaneous Vaginal delivery | 18235 | 91.6 |
| Caesarian Section | 1666 | 8.4 |
| **Place of delivery** |  |  |
| Health facility | 17404 | 87.7 |
| Home/TBA | 2431 | 12.3 |

<: Less than

With regards to umbilical cord care substances, majority of the respondents 16801(85%) did not apply anything to the cord, and only 284 (1%) used chlorhexidine. Out of the 19835 participants, 2752 respondents used non-recommended substances giving an estimated prevalence of 14% (95% CI: 13.40%, 14.35%) among the study participants (**Fig 1)**.

**Fig 1.**
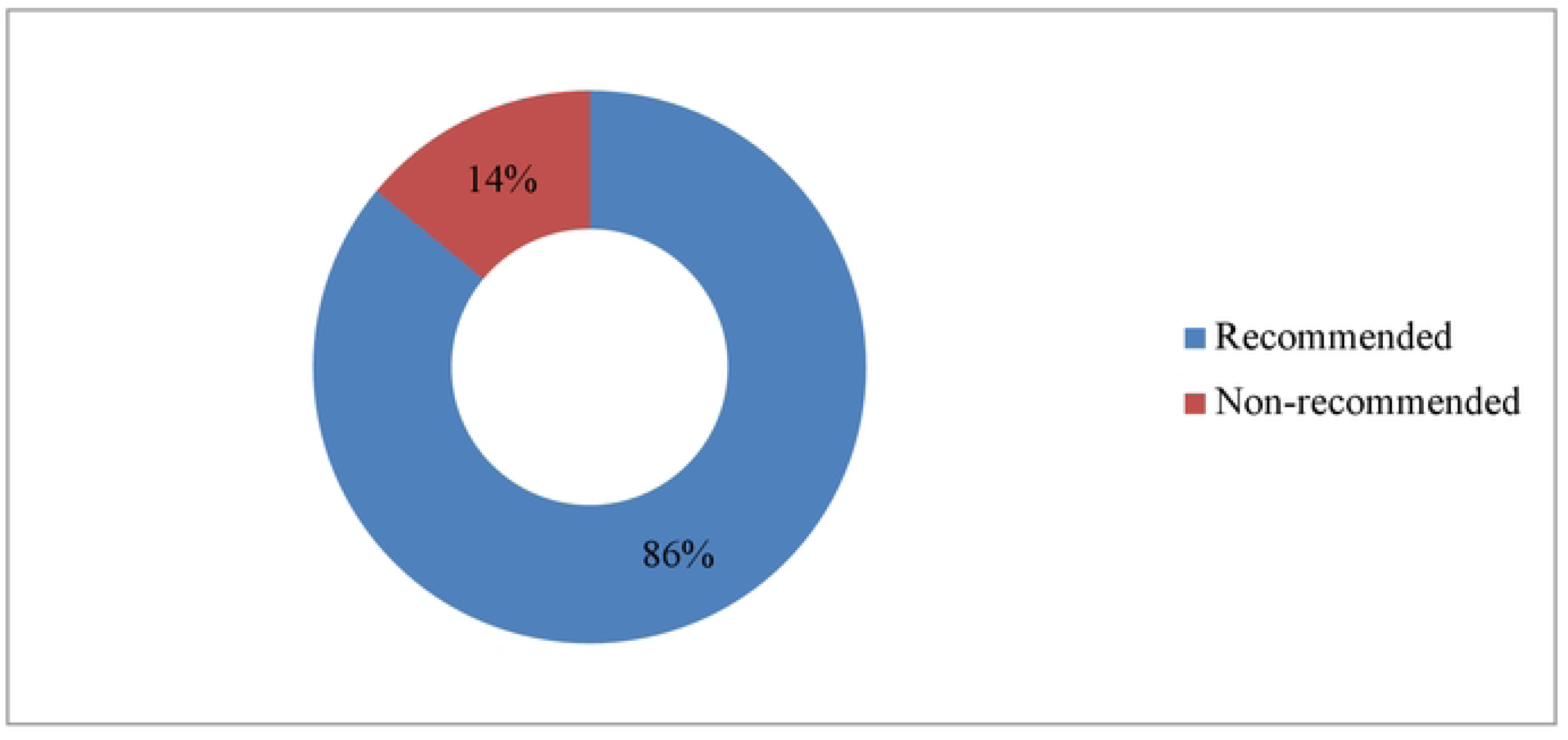
Use of non-recommended substances for Cord care among respondents

At bivariate analysis, key findings revealed that maternal age, religion, PNC attendance and mode of delivery were the factors significantly associated with the use of un-recommended substances for umbilical cord care. Specifically, adolescent mothers were more likely to use non-recommended cord care substances compared to women aged 20-24 years, with a crude prevalence ratio (cPR) of (cPR: 2.74; 95% CI: 2.10-3.58; p<0.001) (table 2). Conversely, older age groups (25-29 and 30+) did not differ significantly from the 20-24 group.

**Table 2.**
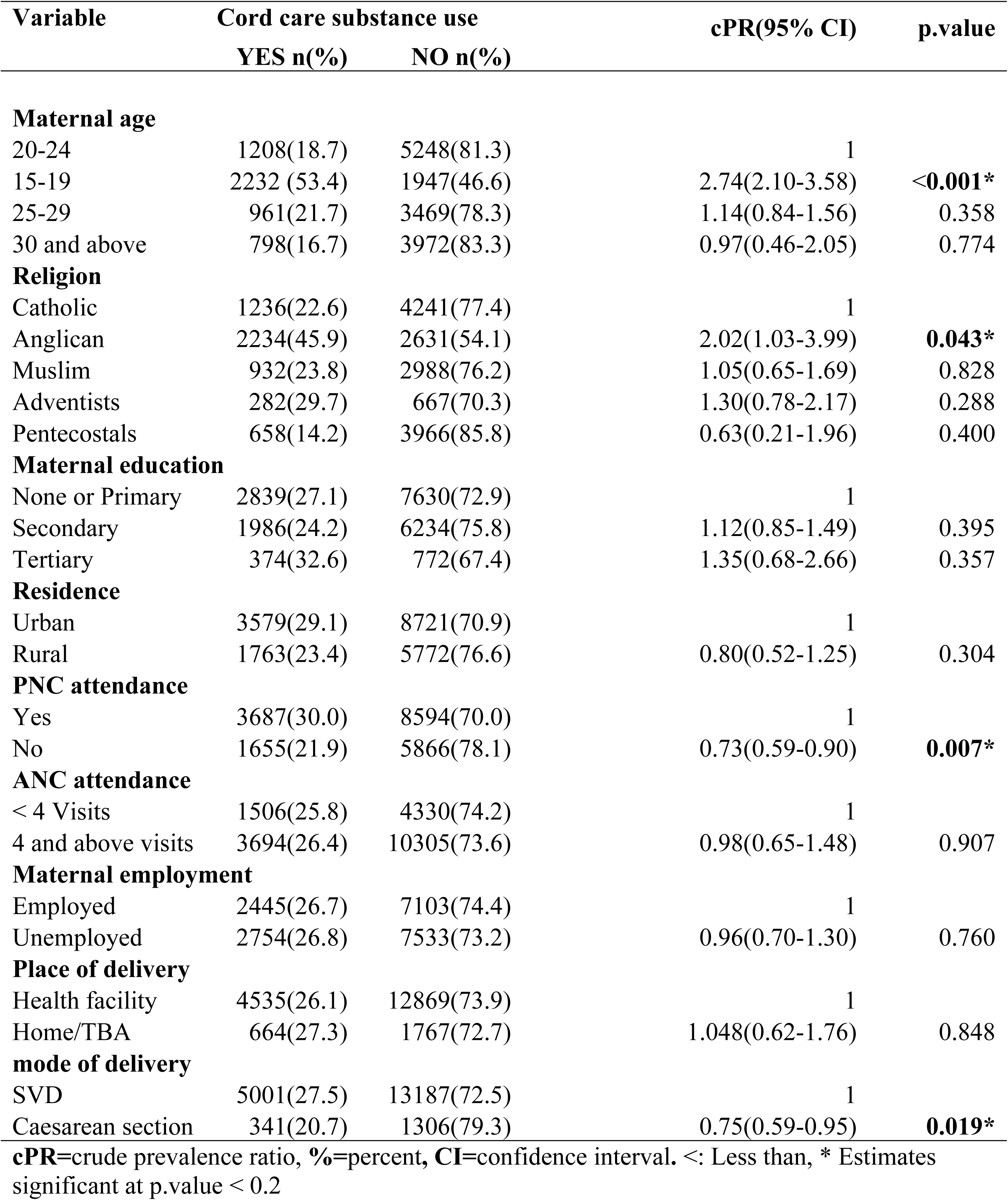
Bivariate analysis for predictors of using non-recommended substances.

Religious affiliation also showed marked differences, Anglican mothers were twice as likely as Catholics to engage in using non-recommended cord care substances (cPR: 2.02; 95% CI: 1.03-3.99; p=0.043), while other religious groups did not show statistically significant differences (table 2)

Importantly, mothers who did not attend PNC were 27% less likely to practice recommended cord care compared to those who did (cPR: 0.73, 95% CI: 0.59-0.90; p=0.007). Furthermore, women who delivered by cesarean section were less likely to use non-recommended substances compared to those who had spontaneous vaginal deliveries (cPR: 0.75; 95% CI: 0.59-0.95, p=0.019). Other factors such as maternal education, residence, ANC attendance, maternal employment, and place of delivery showed no significant association with use of un-recommended cord care substances in this unadjusted analysis **(table 2**).

Religious affiliation, PNC attendance and maternal age remained significant predictor after adjusting for other covariates. Compared to Catholics, mothers who identified as Anglican had 2.25 times higher odds of using non-recommended substances (aPR: 2.25; 95%CI: 1.21-4.18; p=0.014), while Muslim mothers were 48% less likely (aPR: 0.52; 95%CI: 0.38-0.70; P<0.001) to use such substances (table 3). On the same note, Pentecostal mothers were significantly less likely (aPR: 0.15; 95%CI: 0.07–0.34; p < 0.001) to use non-recommended substances for cord care compared to Catholic mothers. Mothers who did not attend postnatal care were 64% less likely to use recommended substances (aPR: 0.36; 95%CI: 0.19-0.67; p=0.004) compared those who attended postnatal care sessions. Teenage mothers, 15-19 years were more than twice as likely to use non-recommended umbilical cord care substances (aPR=2.17; 95% CI:1.1-4.5; p=0.038) compared to mothers in the 20-24 age group (**table 3**).

**Table 3.**
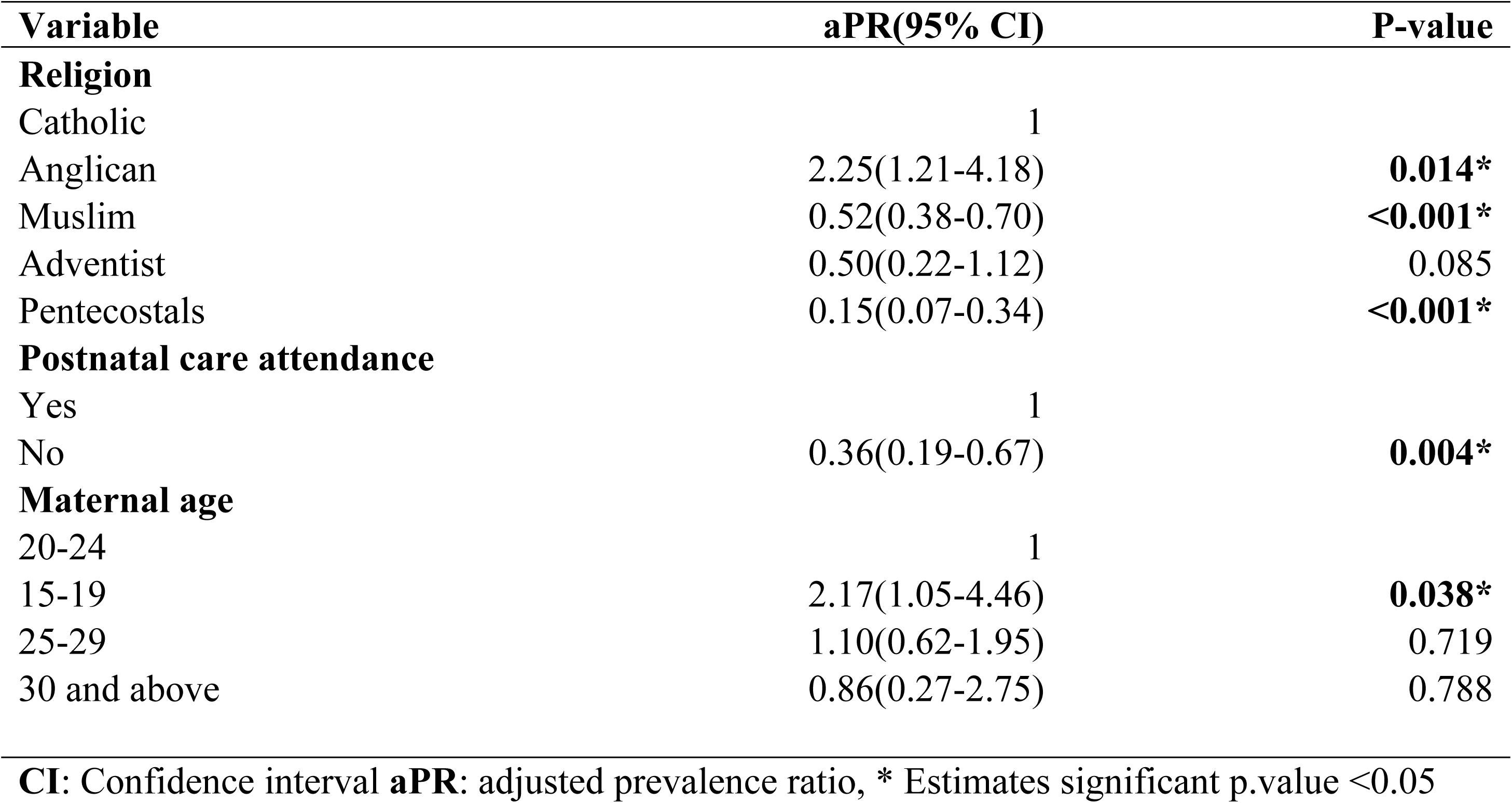
Multivariate analysis for predictors of using non-recommended substances.

| Variable | aPR(95% CI) | P-value |
| --- | --- | --- |
| **Religion** |  |  |
| Catholic | 1 |  |
| Anglican | 2.25(1.21-4.18) | **0.014\*** |
| Muslim | 0.52(0.38-0.70) | **<0.001\*** |
| Adventist | 0.50(0.22-1.12) | 0.085 |
| Pentecostals | 0.15(0.07-0.34) | **<0.001\*** |
| **Postnatal care attendance** |  |  |
| Yes | 1 |  |
| No | 0.36(0.19-0.67) | **0.004\*** |
| **Maternal age** |  |  |
| 20-24 | 1 |  |
| 15-19 | 2.17(1.05-4.46) | **0.038\*** |
| 25-29 | 1.10(0.62-1.95) | 0.719 |
| 30 and above | 0.86(0.27-2.75) | 0.788 |

CI: Confidence interval aPR: adjusted prevalence ratio, \* Estimates significant p.value <0.05

## Discussion

This national household survey provides representative information on the use of non-recommended substances for cord care in Uganda, associated factors and substances commonly used in this context.

### Prevalence and non-recommended substances used for umbilical cord care

In this national study, 14% of mothers, approximately 1 in 10, used non-recommended substances for cord care, thereby exposing newborns to high risk of neonatal sepsis during the most vulnerable newborn period (9, 36).

This prevalence of using non-recommended substances for cord care, 13.9% (CI: 95% CI: 13.40%, 14.35%), was slightly higher than that in studies conducted in Ethiopia (13.7%) and Kenya (12.9%) (8, 37). This high prevalence in Uganda may be attributed to strong cultural beliefs attached to the umbilical cord and the perception that chlorhexidine delays cord separation (17, 20, 23, 38). Such poor cord care practices likely contribute to neonatal infections, deaths and the slow decline of the country’s neonatal mortality rate (39, 40). Additionally, this high prevalence may be due to the absence of specific umbilical cord care recommendations in the EMNCC, and ANC guidelines to educate mothers about acceptable cord care practices (12).

In 2021, the WHO recommended using a 4% chlorhexidine solution or gel in regions with high rates of out-of-facility deliveries, high mortality rates, and dependence on harmful substances. For regions with a high prevalence of health facility deliveries, dry cord care was recommended (11). This shift was based on evidence demonstrating the safety and significant impact of chlorhexidine in reducing neonatal sepsis and mortality (41, 42). Uganda adapted the same strategy but efforts have not be made to make it available to those mostly in need and integrate the recommendation in it’s EMNCC (12).

In this study, non-recommended substances used included; body ointments, oils, baby powder, herbs, saliva, and ash. These substances were similar to those commonly used in many other countries in low, middle and high income settings (17, 43). The similarity in utilization may be attributed to closely share traditional practices and beliefs about healing, infection prevention and bad spirit protection (17, 44). Furthermore, populations in these settings are often faced with under-resourced or inaccessible health care services with variable levels of mistrust in the healthcare systems leaving caregivers with no alternatives other than home-based remedies (45).

### Demographic and Healthcare Utilization Factors

Women 15-19 years were more likely to use non-recommended substances compared to mothers 20-24 years. In contrast, ages 25-29, 30-34, 35-39, and 40-49 years were not significantly associated with this practice. This finding contradicts a study conducted at KIU-TH, where maternal age group 20-24 years was strongly associated with the use of non-recommended substances (9). The higher likelihood of young mothers (15-19 years) engaging in this practice might be attributed to their limited adherence to ANC and PNC visits, and their susceptibility to bad community practices due to limited mothering experience (46). Furthermore, teenage mothers often face increased dependency, peer pressure, and various healthcare system constraints like judgmental healthcare providers, which may deter them from seeking routine hospital-based care (47, 48, 49, 50).

Religion, specifically being Anglican, or Muslim, was strongly associated with use of non-recommended substances for cord care. Although no literature directly associates religion with the use of non-recommended substances for cord care, cultural beliefs related to influence of the cord to the child’s future are documented. For example, in Turkey, the burial place of the cord is believed to determine whether a child will be religious or educated, if buried at mosque, the child will be religious or highly educated if buried at the school gate (51). This suggests that religious and cultural beliefs might indirectly influence various cord care practices.

Not attending postnatal care was significantly associated with the use of non-recommended substances for cord care. Postnatal care is crucial for ensuring good maternal and neonatal outcomes, including proper postnatal practice (52, 53). The findings of this study underscore the importance of PNC attendance to cord care, as those who skipped these visits were more likely to use non-recommended substances. This highlights the need for increased efforts to ensure mothers attend PNC to receive proper guidance on newborn care practices including recommendable cord care.

In this survey, there was no significant association between place of delivery and use of non-recommended substances for cord care. This finding contrasts with a study conducted in Nigeria, which found a strong association between home delivery and the use of non-recommended substances for cord care (54). The discrepancy between these findings could be due to the Nigerian study focusing exclusively on women from rural communities, where home deliveries are more common and healthcare resources may be limited. Additionally, the lack of a significant association in our study might be explained by the fact that majority of mothers delivered from healthcare facilities, which likely guided on proper cord care.

Similarly, there was no significant association between maternal education and the use of non-recommended substances for cord care. This contrasts with several studies that have linked different levels of maternal education with varying cord care practices. For example, some research has shown that having no education is significantly associated with the use of non-recommended care substances (8, 55), while at least primary education linked to beneficial cord care practices (8, 56). The lack of a significant association in our study may be due to the influence of various other factors on umbilical cord care practices, which could overshadow the impact of maternal education alone, and possibly due to the fact that most mothers had at least primary or more education.

Furthermore, the survey found no significant association between antenatal care attendance and the use of non-recommended substances for cord care. This lack of association could be attributed to the fact that majority of the respondents had attended at least one more ANC sessions. Additionally, ANC attendance is generally linked to an increase in mothers’ knowledge regarding recommended newborn care practices, which might have contributed to the overall awareness and reduced variability in cord care practices among the participants (37).

A study by Kalufya documented low knowledge of acceptable cord care practices among rural populations (55). However, contrary to these findings, our study found no significant association between residence (urban vs. rural) and the use of non-recommended substances for cord care. The lack of association might be explained by the universal access to various cord care substances, suggesting that both rural and urban populations have similar levels of exposure and availability, thereby minimizing the impact of residence on cord care practices.

There was no significant association between maternal employment and the use of non-recommended substances for cord care. This finding contradicts a common assumption that individuals in lower socioeconomic status are more likely to use non-recommended substances for cord care (57). However, our findings align with a study conducted in southwest Ethiopia, which also found that a mother’s occupation did not significantly influence cord care practices (58).

This study found no significant association between mode of delivery and the use of non-recommended substances for cord care among mothers who delivered by caesarean section compared to those who delivered by SVD. Given that caesarean sections are typically performed after complications or failed attempts at spontaneous vaginal delivery, mothers of infants born this way may be more vigilant and proactive in their care practices. This heightened level of care could potentially mitigate the differences in cord care practices between the two groups, leading to similar usage rates of non-recommended substances regardless of the delivery mode (59, 60).

This study acknowledge the following limitations; the primary study did not collect data on some variables that were important for this particular study such as parity and cultural practices, limiting our in-depth understanding why mothers chose to use non-recommended substances for cord care. The use of self-reported data about non-recommended substances for cord care might have introduced information and recall bias that might have affected the frequency and quality of data about recommendable or non-recommended substances including the particular substances applied. Recall bias was also a major limitation of this work including possible under reporting/social desirability bias due to the sensitivity of the topic that might have introduced differential or non-differential misclassification. Recall bias was mitigated by abstracting only records for women who had live births in the past 12 months prior to the survey.

### Program and Policy Implications

Given the poor cord care practices and the stagnant neonatal mortality trend, achieving the targets set in the Sharpened Plan II, the National Development Plan IV, and the Health Sector Development Plan’s goal of 19 neonatal deaths per 1000 live births by 2025, as well as the SDG 3.2 target of less than 12 deaths per 1000 live births by 2030, may remain in black and white (27, 31, 61, 62, 63).

The MOH should integrate standardized cord care guidelines into the national EMNCC, recommending 4% chlorhexidine for all mothers given that newborns spend most of their neonatal life in communities with unsure hygiene. Mandate targeted postnatal care messaging on cord care, especially for teenage mothers and women who deliver outside health facilities.

## Conclusions

The prevalence of using non-recommended substances for cord care was high, with up to 1 in every 10 mothers engaging in this practice. The most commonly used substances included body ointments, cooking and motor vehicle oils and herbs. Significant independent factors associated with this practice were maternal age, especially teenage mothers, religion and postnatal care non-attendance.

The MOH, division of health promotion should develop faith inclusive health communication strategies and engage religious leaders as trusted messengers to counter cultural norms that promote use of unsafe cord care substances. It should also design adolescent specific maternal health programs including but not limited to tailored interventions like adolescent mother’s group ANC and PNC. Also, PNC utilization can be strengthened through utilization of community health workers’ follow up, mobile health reminders, and incentives for attendance, as PNC attendance is a key entry point for promoting safe newborn practices.

Further research should explorer; 1) religious and cultural drivers of cord care behaviors in more depth using qualitative approaches to better understand how beliefs influence maternal decision making; 2) test digital or mobile health interventions targeting young and first time mothers with culturally appropriate cord care messages; 2) evaluate the feasibility and cost effectiveness of scaling chlorhexidine distribution through community health systems and faith-based platforms.

## Data Availability

Data is available upon reasonable request to the Principle Investigor of the parent study.

## List of abbreviations

ANC: Antenatal Care
aPR: adjusted prevalence ratios
BCmCH: Bachelors of Clinical Medicine and Community Health
cPR: Crude prevalence ratio
CI: Confidence interval
C.tetani: Clamydia tetani
DE: Design Effect
DHS: Demographic Health Survey
EDHS: Ethiopia Demographic Health Survey Data
EMNCC: Essential Maternal and Newborn Clinical Care Guidelines
FGDs: Focused Group Discussions
HBM: Health Belief Model
KIU: Kampala International University
KIU-TH: Kampala International University Teaching Hospital
MNCH: Maternal Newborn and Child Health
MOH: Ministry Of Health
NM: Neonatal Mortality
NMR: Neonatal Mortality Rate
NPA: National Planning Authority
PNC: Postnatal Care
SDG: Sustainable Development Goal
SOMREC: School Of Medicine Research Ethics Committee
TBAs: Traditional Birth Attendants
TBA: Traditional Birth Attendant
UN: United Nations
UNICEF: United Nations Children’s Fund
UN SDG3: United Sustainable Development Goal 3
UDHS: Uganda Demographic and Health Survey
WHO: World Health Organization

## Declarations

### Ethics approval and consent to participate

Permission to use the data was sought from Makerere University Center of excellence for maternal, newborn and child health and guaranteed by the principal investigator of the primary study.

Permission to do the study was obtained from Makerere University Clinical Epidemiology Unit. Ethical approval was obtained from School of Medicine Research Ethics committee (Mak-SOMREC-2023-824) of Makerere University College of Health Sciences. Data was shared in a de-identified format to protect the confidentiality of the participants.

Data set was accessed from Makerere University Center of Excellence for Maternal, Newborn and Child Health with permission from the primary study principle investigator.

### Consent for publication

A waiver for informed consent and compensation was obtained from SOMREC and consent for publication requirement is not applicable.

### Author’s contributions

NK conceptualized the study. NK, PW, GN, MO RW, RK and DK participated in the review of the protocol.

KN collected and analyzed the data, drafted and revised the draft manuscript.

PW, GN, JN and RK provided expert guidance and revised the manuscript. All authors read and approved the final manuscript.

## Acknowledgement

I would like to extend my heartfelt gratitude to all the Clinical Epidemiology lecturers for imparting invaluable research skills and knowledge throughout my Master’s degree academic journey. A special appreciation goes to Prof. Joan Kalyango for providing an in-depth research experience and unwavering support.

I am profoundly grateful to Prof. Peter Waiswa, my mentor and the principal investigator of the primary study, for his steadfast support and for facilitating the use of the data. Your guidance and insights have been indispensable.

My sincere thanks also go to my supervisors, Dr. Joaniter I. Nankabirwa and Dr. Gertrude Namazzi, for their constructive reviews and guidance throughout the project. Your expertise and advice have significantly contributed to the success of this study.

Lastly, I would like to thank my colleagues, especially Mr. Engole Emmanuel, for constructive criticism and support throughout the program. Your encouragement and collaboration have been greatly appreciated.

